# Contextual and psychological factors of mask-wearing among secondary school students: A cross-sectional survey from Toronto

**DOI:** 10.1101/2023.07.14.23292674

**Authors:** Thomas Liang, Alan Kraguljac, Michelle Science

## Abstract

**Introduction:** After the Ontario government withdrew masking regulations, mask-wearing became a personal choice. Many studies have investigated the factors associated with public mask-wearing, but few have explored the knowledge, attitudes, and psychological factors of masking in high school students. Our study aims to fill this gap.

**Methods:** In February 2023, a cross-sectional survey was distributed online to Grade 9-12 students in a school located in Toronto. Descriptive statistics, correlation analysis, and logistic regression were performed on the quantitative data, while thematic analysis was used to evaluate the qualitative responses.

**Results:** Most of the 62 participants were male with the median age of 16. Approximately half of the respondents reported some frequency of mask-wearing over the past month. Almost all participants claimed to be knowledgeable about COVID-19’s modes of transmission and preventative measures. More participants supported voluntary masking in schools rather than mandated masking. Demographic variables, existing COVID-19 knowledge, and perceived medical benefits were not significantly correlated with masking behaviour. Students who supported voluntary masking were ∼22 times more likely to wear a mask compared to those who held negative attitudes. Participants who felt a high level of perceived barriers were ∼30 times less likely to wear a mask. Mask-wearing individuals cited protection and aesthetic reasons, while the opposition raised arguments about the reduced concern of COVID-19 and downsides of masks.

**Conclusion:** Adolescent mask-wearing is significantly impacted by pre-existing attitudes towards masks, while perceived barriers strongly discourage students from wearing masks. Future research should investigate how to best promote positive beliefs regarding mask-wearing to youth.

## Introduction

During the COVID-19 pandemic, face masking was one of the most important and effective public health measures for slowing the spread of the virus (1–3). After the Ontario government withdrew mandatory mask regulations in June 2022 (4), the choice to mask became a personal decision. However, given the dynamic nature of COVID-19, the individual characteristics and factors of mask-wearing ought to be well-understood to facilitate mass masking if it becomes necessary again. While this topic has been previously investigated among the public (5), few studies have been conducted at the secondary school level. As such, our study aims to fill this gap by evaluating the knowledge, attitudes, and mask-wearing practices (KAP) in high school students. We will also be collecting data on the psychological determinants of masking using the Health Belief Model (HBM) as a guiding framework. Our findings may not only provide insight towards the masking behaviour of adolescents in a mask-optional setting, but also advise for better policies for future respiratory pandemics. Mandatory masking policies in secondary schools have shown to be very effective in slowing the transmission of COVID-19 and its variants (6–10). In Ontario, the masking adherence of children in K-12 schools varied between 43% to 97% across individual studies, where older students generally exhibited higher levels of adherence (11). Gender, age, and grade also proved to be significant factors in influencing masking among students with increased age and grade level correlating with increased mask adherence (12,13). Many studies have found that women were more likely to wear a mask and comply with public mandates (5,14–17), but Guzek et al. (18) reported that male and female individuals were equally likely to wear a mask in the high school context. Female students did, however, show a higher adherence to every other preventative behaviour, such as handwashing and social distancing (18). The level of education and awareness about COVID-19 were also key determinants of mask-wearing behaviour: Secondary school seniors who demonstrated a better understanding of how COVID-19 was transmitted were more likely to wear masks compared to students who claimed that COVID-19 was not deadly (18). Psychological determinants of mask-wearing have not been explored in high school populations, hence the need for this study. Among adults, however, social norms, risk perception, perceived benefits, and social responsibility are most positively associated with mask-wearing tendencies (19–23). Additional motivations include the perceived barriers of masking (e.g. hindrance to communication, difficulty to breathe) as well as relieving fear and anxiety towards COVID-19 (20,22,23).

Interestingly, Wang et al. (20) and Nakayachi et al. (21) reported that the incentive for mask-wearing may actually stem from cosmetic rather than health promotion purposes. Overall, there are many psychological factors that affect an individual’s choice to mask, but the consensus is not clear as to which variables are most influential.

## Methods

### Procedure

We employed a cross-sectional survey to collect primary data on how different contextual and psychological factors affected a high school student’s masking behaviour. After providing their assigned sex and age, respondents were asked to rate their understanding of COVID-19 and its related preventative health measures. They were also asked how often they wore a mask in school, and their position on student masking in school (with and without a government mask mandate). For the psychological determinants, we utilized the HBM and drew inspiration for questions from previous studies.

### Study population and recruitment

The target population for this study was Grade 9-12 high school students, aged 14-18. Due to the resource and time constraints of this project, the sample size was limited to a single, independent school located in Toronto. An open-call email was sent to all eligible students, containing information about the study, participant consent, and the link to the Google Form survey. We also used convenience sampling between different Grade 12 Biology and AP Research classes to obtain a larger sample size.

### Variable measures

To create the variable measures, we drew inspiration or borrowed relevant questions directly from previous studies. We also calculated Cronbach’s alpha coefficients to validate the reliability of multi-item scales (i.e. multiple questions used to evaluate a single variable). A value of >0.6 was considered to be acceptable (24). These variables, along with their corresponding questions and alpha coefficients, are listed in Table 1. Higher mean scores indicated that the participant felt more strongly towards the question prompt.

**Table 1:**
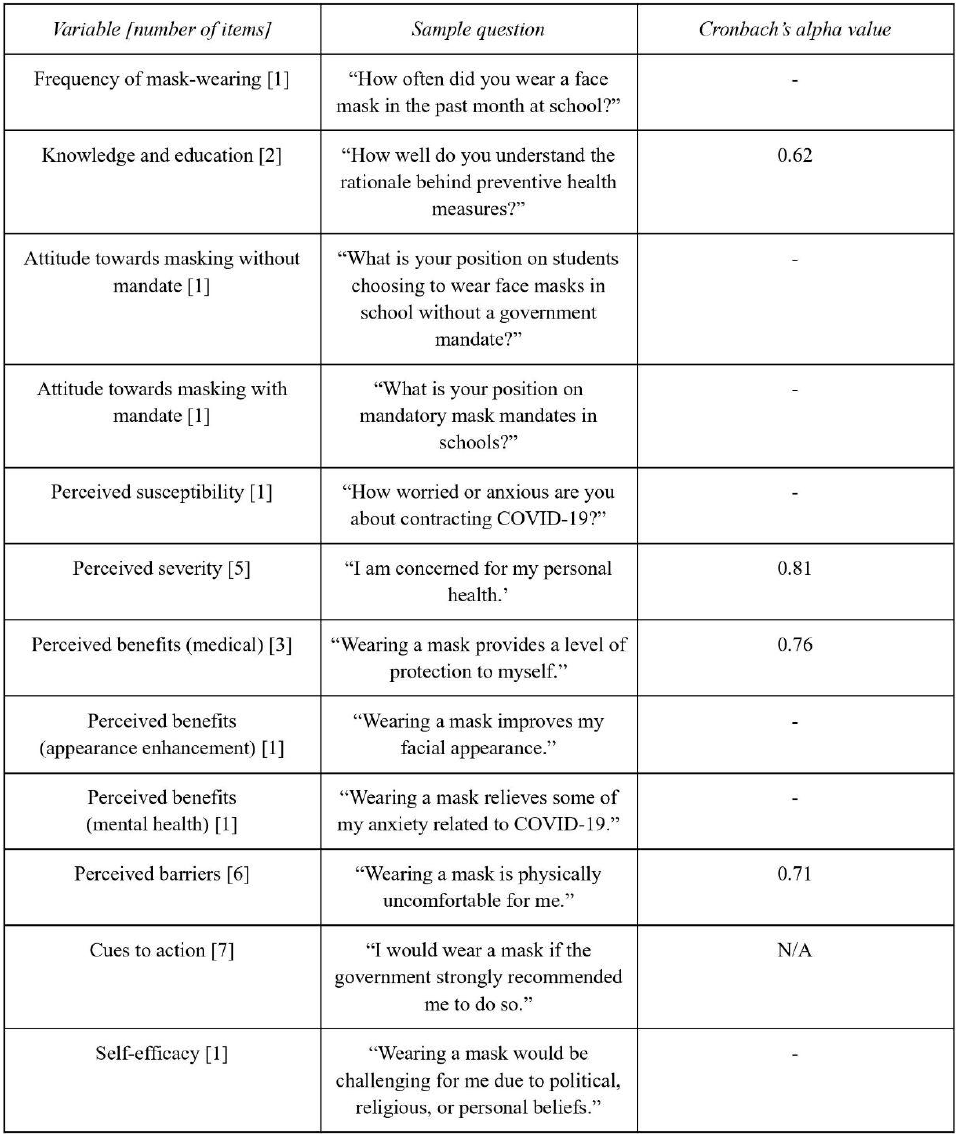
Major study variables of the survey. Sample questions and Crontach’s alpha value for muhr-item scalcs have also been provided.

### Data analysis

All statistical analyses were performed using Microsoft Excel, Version 2304 with Analysis Toolpak Add-in, and IBM SPSS Statistics for Windows, Version 29 (IBM Corp., Armonk, N.Y., USA) (25). Descriptive statistics with counts as percentages were conducted for the KAP data. Because the data on the psychological factors did not follow a normal distribution, Kendall’s tau-b (τ_b_) correlation coefficients were calculated to determine any significant associations between the major study variables. To evaluate the determinants of mask-wearing, logistic regression was performed with “mask usage” as the dichotomous outcome variable and the other contextual and psychological factors as predictors. Odds ratios and 95% confidence intervals were also reported. Statistical significance was determined at the p ≤ 0.05 level for all analyses. Lastly, we used thematic analysis to identify common trends and patterns in the qualitative responses.

### Ethics

This study involved human participants and was approved by the University of Toronto Schools’ Ethics Review Board. Students also provided informed consent prior to starting the survey.

## Results

### Demographic and KAP characteristics of participants

A total of 62 responses were collected for analysis. Most of the participants were male (46.8%) and the median age was 16 (Table 2). Almost half of the participants did not wear a mask at school (48.4%). Most respondents claimed to be “adequately” or “very knowledgeable” (92%) about COVID-19’s modes of transmission, and possessed a “good” to “excellent understanding” (96.8%) of the rationale behind COVID-19 public health measures. In terms of attitudes, approximately half of the participants supported the voluntary usage of face masks in schools without a mandate (54.9%), but the majority were undecided (33.9%) or disapproved (40.3%) of mandatory mask mandates in school.

**Table 2:** Respondents’ demographics. mask-wearing behaviour. COVID-19 knowledge, and attitudes towards masking in schools (N = 62)

| Characteristic | Count, n (%) |
| --- | --- |
| Assigned sex |  |
| Male | 29 (46.8) |
| Female | 27 (43.5) |
| Prefer not to say | 6 (9.7) |
| Age |  |
| 14 | 4 (6.5) |
| 15 | 15 (24.2) |
| 16 | 19 (30.6) |
| 17 | 18 (29) |
| 18 | 6 (9.7) |
| Frequency of mask-wearing at school |  |
| Never | 30 (48.4) |
| Low mask usage | 15 (24.2) |
| High mask usage | 17 (27.4) |
| Understanding of COVID-19's modes of transmission |  |
| Unknowledgeable | 0 (0) |
| Somewhat knowledgeable | 5 (8.1) |
| Adequately knowledgeable | 36 (58.1) |
| Very knowledgeable | 21 (33.9) |
| Understanding of the rationale behind COVID-19 public health measures |  |
| Poor understanding | 0 (0) |
| Mediocre understanding | 2 (3.2) |
| Good understanding | 28 (45.2) |
| Excellent understanding | 32 (51.6) |
| Position on voluntary student masking in school |  |
| Strongly oppose | 2 (3.2) |
| Oppose | 3 (4.8) |
| Neutral | 23 (37.1) |
| Support | 21 (33.9) |
| Strongly support | 13 (21) |
| Position on mandatory mask mandates in school |  |
| Strongly oppose | 7 (11.3) |
| Oppose | 18 (29) |
| Neutral | 21 (33.9) |
| Support | 12 (19.4) |
| Strongly support | 4 (6.5) |

### Correlation analysis

Kendall’s tau-b correlation tests were performed to represent intercorrelations between the 11 study variables (Table 3). There was a moderately positive correlation between masking behaviour and attitude towards masking in schools, both with (τ_b_ = 0.38, p < 0.001) and without a mandate (τ_b_ = 0.49, p < 0.001). Perceived susceptibility, severity, and benefits (appearance enhancement and mental health) were weakly correlated with masking behaviour (τ_b_ = 0.20 - 0.26, p < 0.05), while the correlation between perceived barriers and masking behaviour was negatively correlated (τ_b_ = -0.42, p < 0.001). Interestingly, age, knowledge, and perceived medical benefits were not significantly correlated with mask usage. Lastly, the result of a Chi-square test revealed no statistically significant association between assigned sex and masking behaviour. As expected, attitudes towards voluntary masking were highly correlated with attitudes towards mandated masking (τ_b_ = 0.43, p < 0.001). Perceived susceptibility and severity of the HBM were highly correlated with one another (τ_b_ = 0.54, p < 0.001). Notably, perceived mental health benefits were moderately associated with perceived susceptibility, perceived severity, and perceived appearance-related benefits (τ_b_ = 0.36 - 0.46, p < 0.001).

**Table 3:** Kendall’s tau-b correlations among major study variables.

|  | 1 | 2 | 3 | 4 | 5 | 6 | 7 | 8 | 9 | 10 | 11 | *M* | *SD* |
| --- | --- | --- | --- | --- | --- | --- | --- | --- | --- | --- | --- | --- | --- |
| 1. Masking behaviour | - | -.13 | .03 | .49*** | .38*** | .24* | .20* | .14 | .24* | .26* | -.42*** | 2.15 | 1.37 |
| 2. Age |  | - | .19 | .01 | -.06 | -.05 | -.04 | .10 | -.11 | .02 | .01 | 16.11 | 1.09 |
| 3. Knowledge of COVID-19 |  |  | - | -.01 | .05 | .07 | .10 | .11 | -.02 | .14 | -.06 | 4.37 | .50 |
| 4. Attitude (without mandate) |  |  |  | - | .43*** | .28* | .18 | .25* | .12 | .18 | -.14 | 3.65 | .98 |
| 5. Attitude (with mandate) |  |  |  |  | - | .27* | .42*** | .15 | .33** | .30** | -.21* | 2.81 | 1.08 |
| 6. Perceived susceptibility |  |  |  |  |  | - | .54*** | .17 | .15 | .46*** | .05 | 2.13 | 1.03 |
| 7. Perceived severity |  |  |  |  |  |  | - | .30** | .21* | .40*** | -.04 | 2.94 | .86 |
| 8. Perceived benefits (medical) |  |  |  |  |  |  |  | - | .13 | .33** | -.01 | 4.23 | .58 |
| 9. Perceived benefits (appearance) |  |  |  |  |  |  |  |  | - | .36*** | -.19 | 2.77 | 1.18 |
| 10. Perceived benefits (mental health) |  |  |  |  |  |  |  |  |  | - | -.08 | 2.95 | 1.25 |
| 11. Perceived barriers |  |  |  |  |  |  |  |  |  |  | - | 2.83 | .72 |

The values represent Kendall's tau correlation coefficients, rounded to the nearest hundredth. A correlation coefficient of 0.10 is considered to be a small association; a correlation coefficient of 0.30 is considered a moderate correlation, and a correlation coefficient of 0.50 or larger represents a strong correlation (22). \* $p < 0.05$ , \*\* $p < 0.01$ , \*\*\* $p < 0.001$ . Significant factors are coloured in shades of yellow.

### Regression results

Logistic regression was used to detect which factors were most influential in predicting whether a student was likely to mask or not (Table 4). Participants with more favourable attitudes towards voluntary masking were ∼22 times more likely to wear a mask themselves than those who held less supportive positions (OR = 22.02, 95% CI = 2.78-174.2, p = 0.003). On the other hand, individuals who perceived a high level of barriers to masking were ∼30 times more likely to not wear a mask (OR = 29.87, 95% CI = 2.79-319.47, p = 0.005). All other predictors, despite being significantly correlated in the correlation matrix, were statistically insignificant in the regression model.

**Table 4:** Logistic regression results, including p-values. odds ratios, and 95% confidence intervals.

| Predictor | p-value | Odds ratio (OR) | 95% Confidence Interval (CI) |  |
| --- | --- | --- | --- | --- |
|  |  |  | Lower | Upper |
| Sex | 0.454 | 1.88 | 0.36 | 9.82 |
| Age | 0.773 | 0.89 | 0.39 | 2.02 |
| Knowledge | 0.526 | 0.52 | 0.07 | 4.01 |
| Attitude (without mandate) | 0.003* | 22.02 | 2.78 | 174.20 |
| Attitude (with mandate) | 0.293 | 0.53 | 0.16 | 1.73 |
| Susceptibility | 0.534 | 0.67 | 0.18 | 2.41 |
| Severity | 0.617 | 1.52 | 0.30 | 7.75 |
| Benefits (medical) | 0.410 | 0.39 | 0.04 | 3.65 |
| Benefits (aesthetic) | 0.355 | 0.58 | 0.19 | 1.83 |
| Benefits (anxiety) | 0.101 | 3.12 | 0.80 | 12.18 |
| Barriers | 0.005* | 0.03 | 0.003 | 0.36 |

\*Significant at the $p \leq 0.005$ level.

### Theme analysis of the free-response answers

There were several emergent themes from the qualitative response section (Table 5). Mask-wearing individuals quoted three main motivations: Masking improves their facial appearance, protects themselves and others around them, and is an appropriate response to their anxiety of COVID-19. In contrast, those who chose not to mask raised nine reasons in total, such as the social norm of *not* wearing a mask and the physical discomfort and inconvenience of mask-wearing. Notably, there were two contrasting themes identified in this analysis: the physical appearance aspect of masks and how masks related to one’s anxiety about COVID-19. Some participants viewed mask-wearing as an improvement to their facial appearance (e.g. “Fashion statement”), while another individual considered masks to be a hindrance (e.g. “This dazzling face needs to be out there”). Similarly, one student viewed their moderate anxiety of COVID-19 as a stimulus for personal mask usage (“I couldn’t go into a store or crowded place without one”), while another respondent reported that *not* wearing a mask actually lowered their anxiety associated with the pandemic (“It made them feel like things were ‘normal’ again” and that “wearing a mask brought back some of that worry”).

**Table 5:** Thematic analysis of the participants ’ responses.

| Main Themes | Quotations From Participants |
| --- | --- |
| <u>Reasons for wearing a mask</u> |  |
| 1. Improves facial appearance<sup>1</sup> | "I think I look terrible without a mask honestly."<br>"Fashion statement."<br>"Big nose." |
| 2. Protecting others and oneself | "I'm still masking up to stop the spread to these [vulnerable] populations."<br>"I still wear it [a mask] if I feel possible symptoms of sickness (COVID or otherwise)." |
| 3. Anxiety of COVID-19<sup>2</sup> | "I used to have moderate anxiety about masking in public places (i.e. I couldn't go into a store or crowded place without one), but over the past months have stopped wearing them as frequently at school." |
| <u>Reasons against wearing a mask</u> |  |
| 1. Reduced concern of COVID-19 and the question of necessity | "The dangers are known, but is it really needed is the question as it has become somewhat of a norm."<br>"Even though masks are strongly recommended, I feel that it doesn't seem much of an issue nowadays... This makes me think about whether I need to wear a mask or not."<br>"This is further influenced by the reduced concern surrounding COVID-19 in our communities."<br>"I've been vaccinated 3 times, and have already gotten COVID once, so I'm less worried about reinfection." |
| 2. The social norm of not wearing a mask | "Many students choose not to wear masks in order to fit in with others who do the same."<br>"If I were to show up to school wearing a mask, just knowing my friends, they'd ask why. At least in my friend group, people are used to seeing my full face: It's an understandably (for me) out of the ordinary thing." |
| 3. Taking away from communication | "They take away a fundamental form of communication."<br>"You feel like you're talking to a fucking wall and it's uncomfortable." |
| 4. Impairs facial appearance<sup>1</sup> | "This dazzling face needs to be out there." |
| 5. The feeling of normality<sup>2</sup> | "I am ok to take on the risk of not wearing a mask if it makes me feel like things are "normal" again. Not having to worry about the pandemic nearly as much anymore has made me feel much happier in general, and wearing a mask brings back some of that worry." |
| 6. Inconvenience | "Something I haven't mentioned is simply how much more it [wearing a mask] logistically complicates my life." |
| 7. Minimal benefits | "While the benefits [of wearing a mask] are there, they have been proven to be small/marginal." |
| 8. Physical discomfort | "I also simply feel physically uncomfortable wearing a mask for multiple hours (3+) at a time." |
| 9. Obstructive health conditions | "Adhd moment - I forget mask sometimes so that's is like kind of a problem but oh wellll." |

## Discussion

### Non-significant factors of masking behaviour

Our results found that sex, age, knowledge, and perceived medical benefits did not have a significant correlation or impact on masking behaviour. Historically, prior studies on secondary school students (12) and the public (5,14–17,26) have reported that female and older individuals were more likely to wear masks. Conversely, Rieger (27) and Guzek et al. (28) did not find a relationship between mask usage and gender: This discrepancy may indicate that mask-wearing adolescents do not share the same demographic trends as mask-wearing adults, hinting towards developmental or generational differences. In terms of knowledge, the general trend is that well-educated individuals tend to be more willing to wear masks. However, in instances where the great majority of participants are well-informed about the pandemic (such as the current study), knowledge was not found to be a significant predictor of masking (29,30). Similar to our findings, Wang et al. (20) also concluded that the perceived medical benefits of masks were not a significant predictor of mask-wearing. To the best of our knowledge, no other studies have divided the “perceived benefits” variable into separate components. Collectively, however, perceived benefits have been shown to be an important determinant of masking (16,19,23), but the specific type of “benefit” has not yet been investigated. Our study has, perhaps, shed light on this new area of research.

### What encourages students to mask?

The previous section suggests, then, that mask-wearing individuals choose to do so for different reasons. Most notably, students who supported voluntary masking in schools were ∼22 times more likely to wear a mask themselves, highlighting how pre-existing beliefs, perceptions, or biases about masking may impact one’s masking behaviour (31,32). Likewise, the relationship between perceived susceptibility, severity, and mask usage has been well-established (16,17,20,33,34). It is logical for someone to wear a mask if they feel particularly vulnerable to COVID-19 infection and its consequences (which may especially be true for immunocompromised individuals). For the perceived benefits, however, it is interesting how the participants prioritized the appearance enhancement and mental health benefits of wearing masks over the medical benefits themselves. Some studies have demonstrated how masks tend to increase the perception of facial attractiveness, particularly for those who have conventionally-unattractive faces (35–37), but individuals with higher self-perceived facial attractiveness are actually less likely to wear a mask (38). In a post-COVID era where the health concerns surrounding the pandemic have diminished dramatically, the incentive for mask-wearing may primarily come from its cosmetic purposes instead of self-protection or altruistic motives (38). Regarding mental health, Qin et al. (39) found that frequent mask-wearing had protective associations against psychological distress for schoolchildren. Thus, for students who experience a high degree of anxiety related to the pandemic, masking seems to be an effective tool to lower their symptoms. On the contrary, as identified in the qualitative responses, mask-wearing can also be responsible for bringing back COVID-19 related anxiety. Further research is needed to understand why masks alleviate feelings of anxiety for certain individuals, but aggravate them for others.

### What discourages students from masking?

Perceived barriers presented a significant, negative impact on masking behaviour, meaning that participants who felt greater challenges or difficulties associated with mask-wearing were less likely to wear a mask at school. “Physical discomfort” and “hindrance to communication” were most frequently reported by the participants, which is justified in a high school setting. Because students are expected to engage with their peers and teachers for extensive periods of time, verbal communication with a mask can become irritating for both the speaker and listener due to the sound baffling of the mask as well as the visual omission of the orofacial region (40,41). Furthermore, over the course of the school day, frequent mask-wearing may result in headaches, jaw pain, pain around the ears, and “mask acne”. Another factor that might discourage students from masking regularly is the reduced concern regarding COVID-19 overall. Participants questioned the necessity of wearing masks and expressed how they were not very worried about infection and its consequences. This belief can be attributed to the decreasing number of COVID-19 cases, hospitalizations, and related deaths (45); the increasingly high vaccine coverage (46), and the gradual, societal acceptance of coexisting with the virus. Given the current state of COVID-19, it is highly possible that many students no longer see the *need* to mask.

### Limitations

This investigation had a few limitations. First, the “perceived appearance enhancement” and “mental health benefits” variables were measured on a one-item scale. However, from the discussion, it became clear that the aesthetics and mental health aspects of masking are independent areas of research themselves. Therefore, these one-item measures should serve as broad generalizations, and are not representative of the complexities within the subtopics. Also, since the sample size and scope of our survey were limited, there are restrictions on how applicable our conclusions are to the broader high school population. Future studies should aim to incorporate a larger, more representative sample of students across Ontario.

### Recommendations for policy and future research

To prepare for a future respiratory pandemic in which mass masking becomes necessary again, efforts should be made to improve adolescent attitudes towards masks. As such, the following recommendations are outlined for legislators, public health officials, and researchers. Through public health messaging, the aesthetic benefits of mask-wearing should be emphasized to younger audiences (who tend to be highly impressionable from popular media). For example, seeing their favourite celebrities wearing a mask as a fashion statement (e.g. Alan Walker) may encourage adolescents to don a mask themselves. The government can also incentivize beauty and fashion companies to promote similar masking campaigns. Future research should explore how teenage perceptions of masking can be influenced and improved. However, because support towards masks among Canadian secondary school students is already quite high (47), further studies should instead investigate how to best persuade those with negative attitudes to reconsider (48).

## Data Availability

All data produced in the present study are available upon reasonable request to the authors.

## Acknowledgements

We would like to extend our gratitude to our mentors for their kind advice and guidance during this project: Dr. George Tomlinson and Dr. Alexander Kiss. We would also like to thank the participants for their time in filling out the survey, and the UTS library staff for their assistance in reference management.

